# Mapping performance in 114,237 IVF cycles with donor oocytes reveals reproducible inefficiencies and distinct endophenotypes

**DOI:** 10.1101/2025.09.05.25335105

**Authors:** Christian Simon Ottolini, Francesca Mulas, Rebecca Cavagnola, Elvezia Paraboschi, Ludovica Picchetta, Ruben Ferrando Llobell, María Cruz Palomino, Evelin Lara Molina, Graciela Kohls, Laura Fernández Olmedilla, Maria José De los Santos Molina, Vanessa Vergara Bravo, Antonio Requena Miranda, Antonio Pellicer, Juan Antonio García Velasco, Antonio Capalbo

**Affiliations:** Juno Genetics, Reproductive Genetics, Rome, Italy; Department of Maternal and Fetal Medicine, UCL Institute for Women’s Health, University College London, London, UK; IVIRMA Global Headquaters, Valencia, Spain; IVIRMA Global Research Alliance, Madrid, Spain; IVIRMA Global Research Alliance, Barcelona, Spain; IVIRMA Global Research Alliance, Murcia, Spain; IVIRMA Global Research Alliance, Valencia, Spain; IVI Foundation, Instituto de Investigación Sanitaria La Fe (IIS La Fe), Valencia, Spain; IVIRMA Global Research Alliance, Rome, Italy; Faculty of Medicine, Rey Juan Carlos University, Madrid, Spain; Unit of Molecular Genetics, Center for Advanced Studies and Technology (CAST), “G. d’Annunzio” University of Chieti-Pescara, Chieti, Italy; Department of Psychological Health and Territorial Sciences, School of Medicine and Health Sciences, “G. d’Annunzio” University of Chieti-Pescara, Chieti, Italy

## Abstract

**Background:** In vitro fertilization (IVF) outcomes remain variable and difficult to predict, even in individuals without diagnosed infertility. This uncertainty complicates clinical decision-making and impedes the identification of biological factors that underlie IVF success or failure. A major barrier to progress is the absence of standardized criteria to distinguish true biological inefficiencies from random variation or technical noise in IVF procedures.

**Objective:** To develop a robust statistical framework for defining IVF under- and overperformance in a fertile population, enabling precise phenotyping of IVF inefficiencies and laying the foundation for future genomic and mechanistic studies.

**Methods:** We conducted the largest retrospective cohort analysis to date of IVF outcomes in 114.237 oocyte donation cycles, from 32.492 rigorously screened donors who underwent 84.228 stimulation cycles. IVF performance was evaluated across four developmental stages: oocyte maturation, fertilization, blastocyst formation, and preimplantation development efficiency. Donors were classified as statistical under- or overperformers using a combination of binomial testing and mixed-effects logistic regression models, adjusting for identified confounders. Reproducibility of performance classifications was assessed through multi-cycle consistency analyses and supervised predictive modelling.

**Results:** Despite stringent fertility screening, 0.9% to 2.1% of donors were identified as statistically underperformers in individual developmental metrics, with similar proportions observed for overperformers. Minimal overlap between donors falling into significant categories across different developmental metrics supported the interpretation of IVF performance as comprising biologically distinct endophenotypes. Underperformance was reproducible across multiple cycles in a subset of donors (P < 0.001), indicating a likely biological basis rather than random or procedural variability. Logistic regression models demonstrated modest predictive ability for future outlier status (AUC > 0.61 and < 0.71), further supporting the presence of stable, donor-specific traits. Interestingly, comparison of donor clinical variables revealed a possibly clinically meaningful difference in the distribution of donors that had a documented live birth prior to donation between performance groups for all developmental metrics, indicating a potential association with proven fertility and better IVF outcomes. Finally, 109 donors across performance groups were surveyed, revealing no identifiable differences in natural fertility outcomes, further reinforcing that IVF inefficiency does not necessarily reflect underlying infertility.

**Conclusion:** This unprecedented and methodologically rigorous analysis demonstrates that statistically significant deviations in IVF outcomes can occur even among presumed fertile donors. These findings challenge the assumption that poor IVF outcomes are inherently indicative of infertility and support a shift toward defining IVF failure through biological, rather than purely clinical, criteria. Our standardized phenotyping framework offers a powerful tool for identifying the molecular and genetic contributors to IVF inefficiency and for advancing personalized reproductive medicine.

## Introduction

Recognized by the World Health Organization as a disease of the male or female reproductive system, infertility affects millions worldwide, with its prevalence increasing due to factors such as delayed childbearing, lifestyle changes, and environmental exposures^1^. While anatomical anomalies can often be diagnosed, it is estimated that around half of infertility cases are directly linked to genetic defects^2^ with only a small fraction currently receiving a definitive molecular diagnosis^3^.

In vitro fertilization (IVF) has revolutionized reproductive medicine, offering a pathway to parenthood for individuals and couples suffering from infertility who are unable to conceive naturally. However, IVF outcomes remain unpredictable, even in individuals without infertility diagnoses or with proven fertility, including those undergoing preimplantation genetic testing for monogenic disorders^4,5^. Poor IVF outcomes are often presumed to reflect an underlying fertility issue, yet this assumption overlooks cases where otherwise fertile individuals experience unexpectedly poor IVF performance. Distinguishing true infertility from IVF-specific inefficiencies, or indeed random variations, is essential for refining prognostic models, optimizing treatment strategies, and deepening our understanding of reproductive physiology.

Advancements in molecular genetics have begun to shed light on the genetic factors underlying infertility and IVF failure^6^. Pathogenic variants affecting gamete maturation, fertilization, and early embryo development have been associated with poor IVF outcomes^5,7–11^. However, genetic discovery studies in this field have primarily focused on monogenic causes of absolute IVF failure, often in consanguineous populations where homozygous pathogenic variants are more readily identified, and generally they have included only a handful number of families studied for targeted gene-panels. While these studies provide valuable insights into reproductive biology, they frequently lack generalizability and fail to capture the broader genetic heterogeneity of infertility and among populations of different ancestry. Moreover, by concentrating on extreme infertility phenotypes, they overlook subtler genetic contributions that may influence fertility treatment outcomes without causing absolute infertility or IVF failure.

As our understanding of reproductive genetics advances, so too does the need for integrating comprehensive testing strategies into ART workflows^3,12^. A critical challenge in developing such tools in reproductive medicine is the absence of a standardized framework for identifying individuals who consistently experience poor IVF outcomes. Without clear criteria, it remains difficult to distinguish between random variation and intrinsic inefficiencies in IVF procedures. This limitation hampers the development of accurate predictive models, complicates patient counselling, and impedes the personalization of assisted reproductive treatments. Furthermore, the lack of a systematic approach to defining IVF failure slows progress in uncovering the biological mechanisms underlying poor outcomes. Establishing robust methodologies to phenotype IVF inefficiency is therefore crucial for advancing research, improving clinical decision-making, and developing targeted interventions to mitigate these challenges.

A fundamental step in this effort is defining a baseline for underperforming IVF outcomes, establishing a reference point against which poor outcomes can be objectively assessed. Without an established baseline, it remains challenging to differentiate technical variation from true biological IVF outliers. Understanding the extent to which IVF underperformance occurs in fertile individuals would enable researchers and clinicians to quantify the inefficiencies inherent to IVF, determine whether poor outcomes fall within a normal statistical range, and systematically investigate potential underlying causes. Such a framework is essential not only for improving the interpretation of individual IVF failures but also for identifying broader trends linked to biological, technical, or procedural factors^3^. Ultimately, characterizing IVF outliers will provide the foundation for association studies aimed at identifying genetic, epigenetic, and environmental contributors to IVF inefficiency, paving the way for improved diagnostics and interventions.

To address this gap, we conducted a statistical analysis of unprecedented scale on IVF outcomes from oocyte donation cycles. Oocyte donors provide an ideal model for studying IVF performance independently of infertility, as they undergo rigorous fertility screening and often demonstrate proven fertility. By leveraging our unprecedentedly large dataset of oocyte donors, we assessed the extent to which even a highly screened, presumed fertile population can exhibit unexpectedly poor IVF outcomes. To further support these findings, we conducted a survey among underperforming donors, collecting self-reported data on their subsequent natural fertility, including conception history, time to pregnancy, and reproductive success. This additional layer of evidence reinforces the notion that statistically poor IVF outcomes can arise even in truly fertile individuals, highlighting the need for a standardized framework to identify and address IVF inefficiencies.

## Results

### Donor demographics

The dataset included a total of 32,492 egg donors, 84,228 stimulation cycles, and 2,413,697 retrieved oocytes, ultimately resulting in 114,237 donation cycles involving 85,999 oocyte recipients (Figure 1 A). Baseline donor characteristics are summarized in Figure 1 and Table 1.

**Figure 1.**
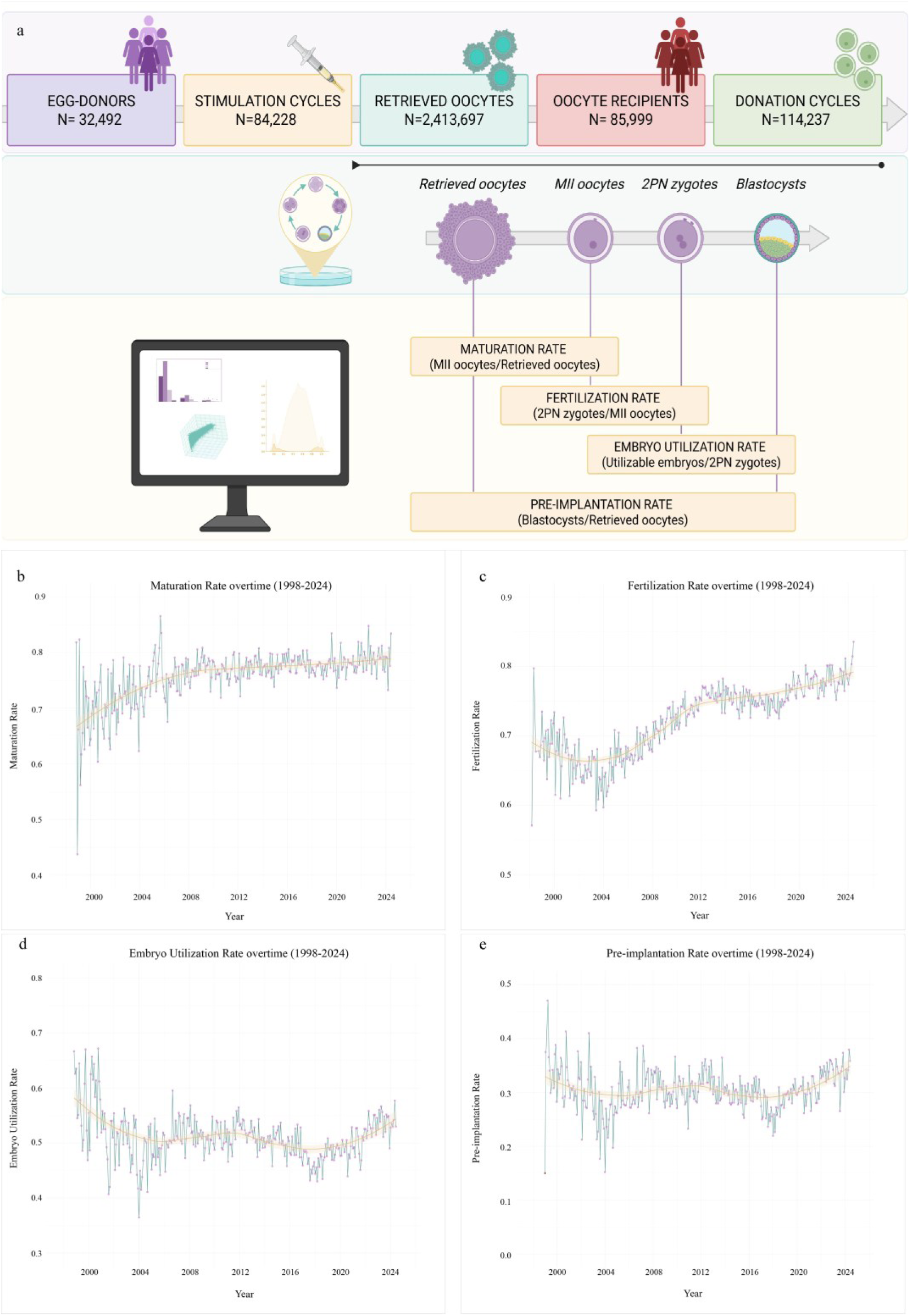
Structure of the dataset and temporal trends in IVF developmental outcomes. (a) Overview of the dataset composition, including total number of oocyte donors, stimulation cycles, and IVF donation cycles analyzed. (b–e) Temporal trends in four key IVF developmental rates from 1999 to 2023: (b) oocyte maturation rate, (c) fertilization rate, (d) blastocyst formation rate, and (e) preimplantation development efficiency.

**Table 1.** Summary of the number of donors and cycles included in each developmental rate analysis after quality control (QC), along with key donor and cycle characteristics. The table compares these QC subsets to the full dataset to check for any differences that might indicate selection bias. P-values show whether there were statistically significant differences between the QC subsets and the full dataset. Effect sizes quantify how large those differences were, as shown by Magnitude levels “negl.” means negligible (likely not meaningful), “n/a” means not applicable (the difference was not statistically significant, so no effect size was calculated).

| Variable | Category | Mean ± SD / Percentage |  |  |  |  | Comparisons vs Global |  |  |  |  |  |  |  |  |  |  |  |
| --- | --- | --- | --- | --- | --- | --- | --- | --- | --- | --- | --- | --- | --- | --- | --- | --- | --- | --- |
|  |  | Global | Mat. | Fert. | Emb-Util. | Pre-implant. | Maturation |  |  | Fertilization |  |  | Embryo-Utilization |  |  | Pre-implant. |  |  |
|  |  | 32,492 | 27,469 | 32,342 | 31,263 | 14,408 | P-val | Eff. size | Magn. | P-val | Eff. size | Magn. | P-val | Eff. size | Magn. | P-val | Eff. size | Magn. |
| Oocytes retrieved | — | — | 18.0±8.2 | 18.8±8.1 | — | 17.7±7.5 | — | — | — | — | — | — | — | — | — | — | — | — |
| Fertilized oocytes 2PN | — | — | — | 7.8±2.5 | 7.9±2.5 | — | — | — | — | — | — | — | — | — | — | — | — | — |
| Oocytes MII | — | — | 13.9±6.8 | — | — | — | — | — | — | — | — | — | — | — | — | — | — | — |
| Usable blastocyst | — | — | — | — | 4.0±2.1 | 5.2±3.4 | — | — | — | — | — | — | — | — | — | — | — | — |
| Age (years) | — | 25.3±4.3 | 25.1±4.3 | 25.2±4.3 | 25.2±4.3 | 25.1±4.3 | 0.081 | n/a | n/a | 1.000 | n/a | n/a | 0.527 | n/a | n/a | 1.000 | n/a | n/a |
| BMI | — | 22.6±3.0 | 22.6±3.0 | 22.6±3.0 | 23.9±3.1 | 24.6±3.0 | 1.000 | n/a | n/a | 1.000 | n/a | n/a | 0.158 | n/a | n/a | 0.301 | n/a | n/a |
| Stimulation dose | — | 2361±647 | 2350±630 | 2362±647 | 2363±646 | 2350±640 | 0.712 | n/a | n/a | 1.000 | n/a | n/a | 0.135 | n/a | n/a | <0.001 | -0.16 | negl. |
| Ethnic background | White | 97.4% | 96.8% | 97.4% | 97.4% | 97.8% | — | — | — | — | — | — | — | — | — | — | — | — |
|  | Black | 2.4% | 2.9% | 2.4% | 2.4% | 1.9% | <0.001 | 0.01 | negl. | 0.99 | n/a | n/a | 0.97 | n/a | n/a | 0.015 | n/a | n/a |
|  | Asian | 0.3% | 0.3% | 0.3% | 0.3% | 0.0% | — | — | — | — | — | — | — | — | — | — | — | — |
| Type of trigger | hCG | 53.1% | 48.9% | 48.9% | 50.1% | 52.4% | — | — | — | — | — | — | — | — | — | — | — | — |
|  | GnRH-a | 46.7% | 51.0% | 51.0% | 49.2% | 47.5% | <0.001 | 0.04 | negl. | <0.001 | 0.02 | negl. | <0.001 | 0.09 | negl. | <0.001 | 0.02 | negl. |
|  | Other | 0.2% | 0.1% | 0.1% | 0.2% | 0.0% | — | — | — | — | — | — | — | — | — | — | — | — |

Temporal trends in IVF performance across the four developmental rates over the study period demonstrated consistent improvements, reflecting advances in both clinical and laboratory practices (Figure 1B–E). Maturation and fertilization rates increased steadily over time, consistent with observed changes in the use of different stimulation regimen. The embryo utilisation rate showed a non-linear pattern, with an initial decline followed by a gradual improvement in more recent years, likely due to shifts and improvements in embryo culture. Similarly, the pre-implantation rate exhibited a gradual upward trend, suggesting marked improvements in the overall efficiency of the IVF process in the oocyte donation programme over time. These temporal changes were systematically measured and statistically controlled for in downstream analyses to ensure accurate comparisons across clinics and protocols.

Following quality control procedures (i.e., removal of incomplete or inconsistent records), 27.469, 32.342, 31.263, 14.408 donors and 68.966, 113.351, 107.046, 27.404 oocyte donation cycles remained in the final subsets for analysis of maturation, fertilisation, embryo utilisation, and preimplantation development rates respectively (Table 1).

The demographic characteristics remained broadly consistent across donors post-QC and among those classified under each developmental outcome rate with negligible effects observed compared to the full dataset, suggesting that no selection bias was introduced during quality control or subgroup classification (Table 1).

An exploratory analysis was then conducted to evaluate whether fertilisation and embryo developmental outcomes were influenced by characteristics of the sperm sample used by the recipient. Available data included sperm origin (i.e. partner, donated or both), sperm source (i.e., ejaculated, epididymal aspiration, or testicular biopsy) and the sperm concentration of the ejaculate used for insemination. No significant associations were observed with fertilisation and embryo utilisation rates (p<0.05). Though a significant association was found for the Preimplantation rate (P=0.000376), the calculated Beta effect size was negligible (B=0.0003523), indicating a clinically unmeasurable impact (Supplementary table 1).

### Distribution of developmental outcomes and identification of statistical over and under performers

To assess variability in IVF performance across the donor population, we generated probability density plots for four key developmental metrics. These plots illustrate the distribution and shape of performance outcomes across all cycles and donors (Figure 2, A, B, C, D). The maturation and fertilization rates were notably skewed towards higher values, indicating an overall high average value of the rates (0.77±0.184 and 0.74±0.152, respectively) in the population. In contrast, preimplantation development efficiency exhibited a shift towards lower values (0.30±0.164). Embryo utilization rate demonstrated a broader, more symmetric distribution (0.50±0.221), reflecting greater normal variability. These distributions influenced the classification of under- and over-performing donors, where phenotypes with a skewed distribution resulted in a wider range of rates observed for under- and over-performing donors.

**Figure 2.**
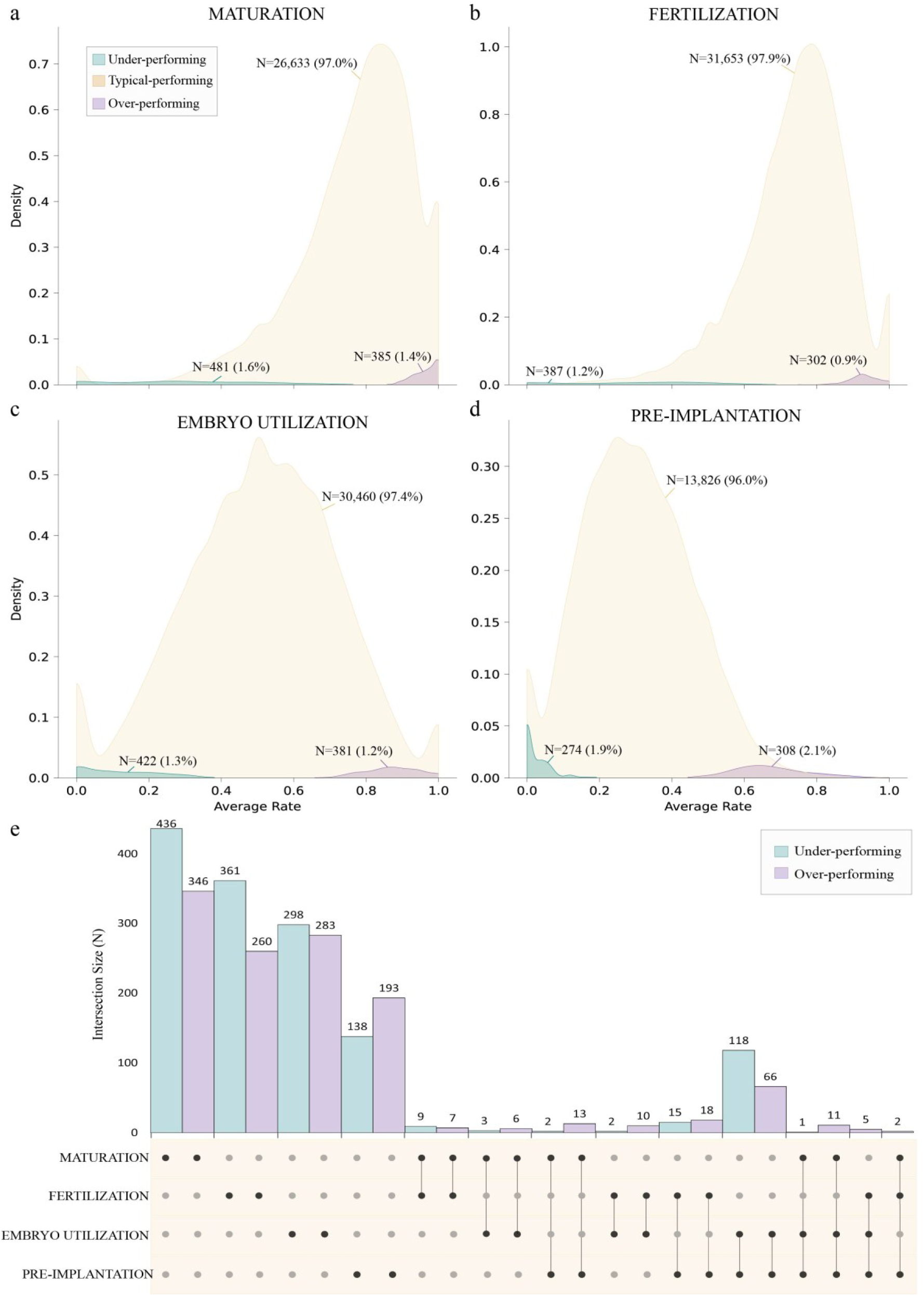
Distribution of donor performance across four key IVF developmental metrics. Probability density plots illustrate the distribution of donor performance for (a) oocyte maturation rate, (b) fertilization rate, (c) embryo utilization rate, and (d) preimplantation development efficiency. Number and percentage of donors classified as underperforming, typical and overperforming are indicated in each density plot. (e) Overlap of statistical outliers across developmental phenotypes.

The number and proportion of under- and overperforming donors varied across the four developmental phenotypes (Figure 2, A, B, C, D). Underperformance was lowest at fertilization (1.2%) and embryo utilization (1.3%), and highest at maturation (1.6%), and pre-implantation development (1.9%). A similar pattern was observed for overperformance, which ranged from 0.9% for fertilization to 2.1% for pre-implantation rate. These proportions reflect both the intrinsic variability of each biological process and the distribution shape of each outcome metric.

We next evaluated whether under or overperformance in one developmental stage was predictive of similar patterns in others. Overlap between classification categories was minimal for both underperformers (Jaccard_avg: 0.042) and overperformers (Jaccard_avg: 0.046) (Figure 2, E), suggesting that poor or exceptional outcomes in one metric did not strongly predict deviation in another, supporting the interpretation of these IVF metrics as distinct biological endophenotypes, with largely non-overlapping performance profiles.

### Characteristics of over- and underperforming donors

Available demographic and clinical characteristics were compared across underperformers, overperformers, and typical performers (Table 2). Post-hoc analyses revealed significant differences across the four phenotype pairings, particularly in donor age, stimulation dose, trigger type, and proven fertility (defined as documentation of a live birth prior to donation). However, with the exception of proven fertility, all comparisons showed small or negligible effect sizes, suggesting that observed differences may reflect sample imbalance or random variation rather than clinically meaningful distinctions. Donors with proven fertility were significantly more prevalent in the overperforming group compared to the underperforming group across all four developmental rates, with a medium effect size (Table 2). This suggests a modestly increased likelihood of overperforming outcomes among donors with prior live births, and a higher likelihood of underperforming outcomes among those without proven fertility.

**Table 2.** Comparison of key donor and cycle characteristics across performance groups (underperformers, typical performers, and overperformers) for each of the four IVF developmental rates. This table assesses whether demographic or clinical variables are associated with classification as a statistical over- or underperformer. Pairwise comparisons between performance groups were conducted to evaluate potential selection biases or confounding factors that might explain differences in IVF outcomes. P-values show whether there were statistically significant differences between the QC subsets and the full dataset. Effect sizes quantify how large those differences were, as indicated by Magnitude levels: “negl.” (negligible, likely not meaningful), “small” (small, likely not meaningful), “med” (medium, could reflect a biologically or clinically relevant trend, even if not strong enough to serve as a standalone predictive marker) and “n/a” (not applicable: the difference was not statistically significant, so no effect size was calculated).

| Variable | Test | Maturation |  |  | Fertilization |  |  | Embryo-Utilization |  |  | Pre-implantation |  |  |
| --- | --- | --- | --- | --- | --- | --- | --- | --- | --- | --- | --- | --- | --- |
|  |  | P-value | Eff. size | Magn. | P-value | Eff. size | Magn. | P-value | Eff. size | Magn. | P-value | Eff. size | Magn. |
| Age | Kruskal-Wallis | <0.001 | 0.001 | negl. | <0.001 | 0.000 | negl. | <0.001 | 0.001 | negl. | <0.001 | 0.004 | negl. |
|  | Under-Over | <0.001 | -0.173 | negl. | 0.002 | -0.149 | negl. | <0.001 | -0.167 | negl. | <0.001 | -0.331 | small |
|  | Under-Typical | 0.207 | n/a | n/a | 0.729 | n/a | n/a | 0.118 | n/a | n/a | <0.001 | -0.234 | small |
|  | Over-Typical | <0.001 | 0.136 | negl. | <0.001 | 0.135 | negl. | <0.001 | 0.125 | negl. | 0.005 | 0.093 | negl. |
| BMI | Kruskal-Wallis | 0.477 | n/a | n/a | 0.831 | n/a | n/a | 0.349 | n/a | n/a | 0.376 | n/a | n/a |
| Stimulation dose | Kruskal-Wallis | 0.001 | 0.001 | negl. | 0.062 | n/a | n/a | 0.001 | 0.000 | negl. | 0.195 | n/a | n/a |
|  | Under-Over | 0.357 | n/a | n/a | n/a | n/a | n/a | 0.391 | n/a | n/a | n/a | n/a | n/a |
|  | Under-Typical | 0.005 | -0.13 | negl. | n/a | n/a | n/a | 0.076 | n/a | n/a | n/a | n/a | n/a |
|  | Over-Typical | 0.113 | n/a | n/a | n/a | n/a | n/a | 0.005 | -0.094 | negl. | n/a | n/a | n/a |
| Ethnic background | Chi-square | 0.026 | n/a | n/a | 0.558 | n/a | n/a | 0.015 | n/a | n/a | 0.387 | n/a | n/a |
| Type of triggering | Chi-square | <0.001 | 0.028 | negl. | <0.001 | 0.053 | negl. | <0.001 | 0.025 | negl. | 0.096 | n/a | n/a |
|  | Under-Over | 0.818 | n/a | n/a | <0.001 | 0.193 | small | <0.001 | 0.142 | small | n/a | n/a | n/a |
|  | Under-Typical | <0.001 | 0.025 | negl. | <0.001 | 0.052 | negl. | <0.001 | 0.022 | negl. | n/a | n/a | n/a |
|  | Over-Typical | 0.087 | n/a | n/a | 0.153 | n/a | n/a | 0.267 | n/a | n/a | n/a | n/a | n/a |
| Proven Fertility | Chi-square | <0.001 | 0.074 | negl. | <0.001 | 0.064 | negl. | <0.001 | 0.055 | negl. | <0.001 | 0.119 | small |
|  | Under-Over | <0.001 | 0.418 | med. | <0.001 | 0.398 | med. | <0.001 | 0.343 | med. | <0.001 | 0.495 | med. |
|  | Under-Typical | <0.001 | 0.05 | negl. | <0.001 | 0.051 | negl. | <0.001 | 0.037 | negl. | <0.001 | 0.109 | small |
|  | Over-Typical | <0.001 | 0.053 | negl. | <0.001 | 0.037 | negl. | <0.001 | 0.040 | negl. | <0.001 | 0.045 | negl. |

Over and underperformance was shown also to be reproducible across cycles. Among donors with three or more stimulations, consistent classification as under- or overperformers occurred significantly more often than expected by chance (P < 0.001; Fig. 3A). As expected, the variance of performance estimates declined steadily as additional cycles were aggregated, reflecting improved precision with larger sample sizes (Fig. 3B). Furthermore, a logistic regression classification model was used to predict statistical over- or underperformance in subsequent stimulation cycles across the four key developmental rates. Model performance was evaluated using receiver operating characteristic (ROC) curves, as shown in Figure 3C and 3D for underperformers and overperformers, respectively. The model demonstrated consistent predictive ability, with area under the curve (AUC) values ranging from 0.61 to 0.71 for underperformance across all four developmental rates. For overperformance, AUC values ranged from 0.61 to 0.69 for maturation, fertilization, and embryo utilization rates. Although most AUC values fell below the threshold typically considered sufficient for clinical application (AUC ≥ 0.75), these results together nonetheless reflect a reproducible biological signal and support the presence of stable, donor-specific inefficiencies in IVF performance across cycles.

**Figure 3.**
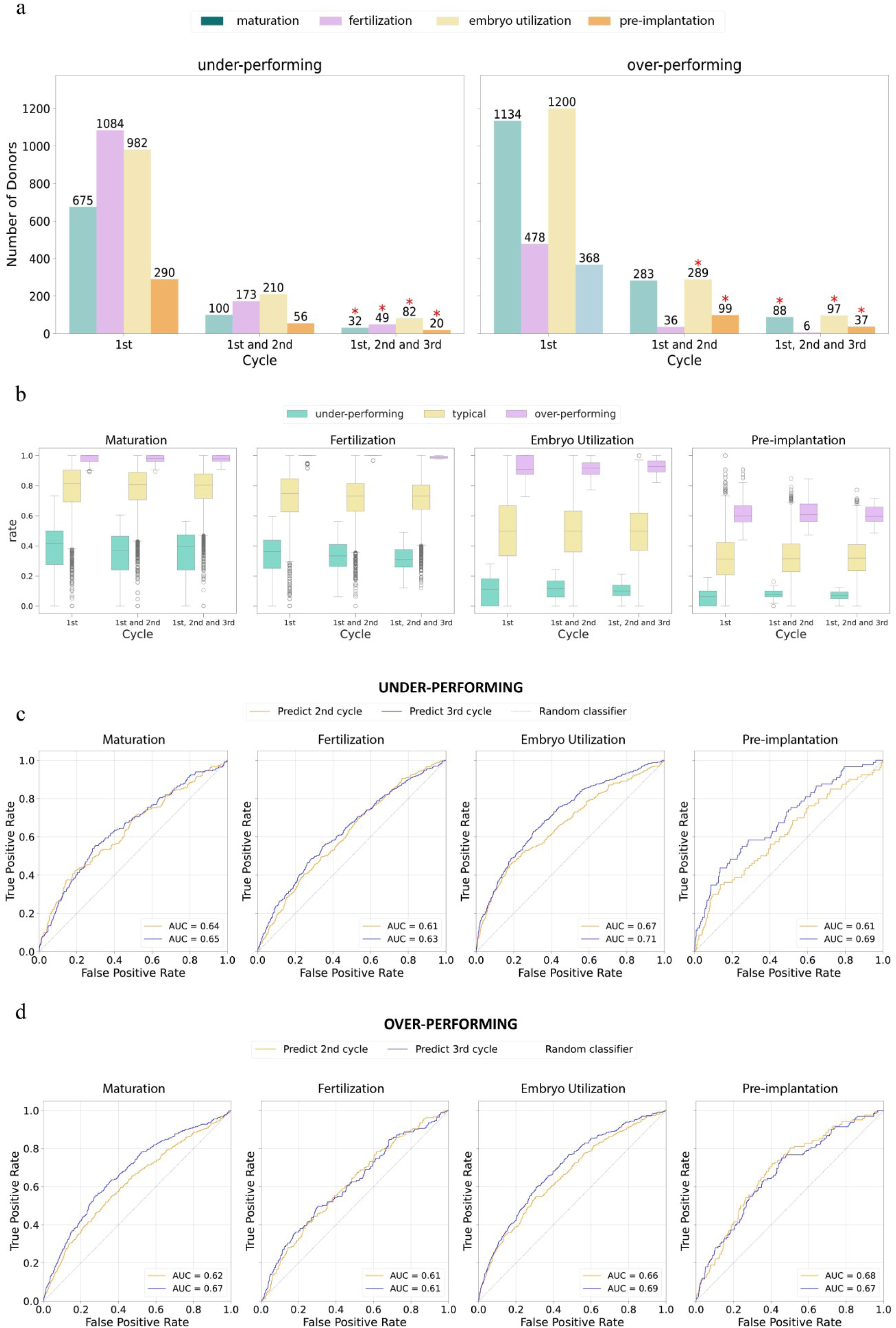
Reproducibility and predictability of donor IVF performance across multiple cycles. (a) Schematic overview of the predictive framework and bar charts showing the proportion of donors with three or more stimulation cycles who were consistently classified as statistical underperformers or overperformers, respectively, in the same developmental rate. * Indicates a statistically significant enrichment in the observed frequency of consistent classifications compared to what would be expected by chance alone (P < 0.001), supporting the biological reproducibility of performance patterns. (b) Box plots showing the distribution of developmental outcomes (rate values) for donors classified as underperformers, typical performers, or overperformers across the four IVF developmental rates. Results are cumulative across one, two, or all available cycles per donor. (c and d) Receiver Operating Characteristic (ROC) curves for logistic regression models predicting underperformance and overperformance, respectively, using data from the first cycle alone or from the first and second cycles combined.

### Post donation survey results

A total of 109 donors completed the structured questionnaire, including donors classified as typical, underperformers and overperformers for each developmental rate: 100, 4 and 5 for Maturation, 98 and 11 (no overperformers) for Fertilization, 87, 14 and 8 for Embryo utilization and 93,8 and 8 for Pre-implantation, respectively. Across all surveyed donors, there were no statistically significant differences in self-reported natural fertility outcomes, such as time to pregnancy, number of natural pregnancies, or pregnancy losses, between the three performance groups (Supplementary table 2). Within the subset of embryo utilization and preimplantation development outliers for whom IVF live birth outcomes from donation cycles were available, no significant associations were found with live birth and any individual questionnaire responses (Supplementary table 3). However, there was a trend suggesting that donors who reported taking longer than one year to conceive naturally, or who did not conceive despite trying, appeared less likely to have had a live birth from their donated oocytes (Supplementary figure 1). While this trend did not reach statistical significance, it may reflect a subtle biological association that warrants further investigation in a larger respondent cohort.

## Discussion

This study provides the largest retrospective analysis to date of IVF outcomes among oocyte donors, a population typically presumed to have optimal fertility. By analysing over 110.000 IVF cycles from more than 32.000 donors, we demonstrate that statistically significant underperformance can occur across key developmental stages even in individuals with no apparent infertility.

Our analysis defined four developmental phenotypes: (1) oocyte maturation, (2) fertilization, (3) embryo utilisation, and (4) overall preimplantation efficiency, identifying distinct groups of statistical underperformers representing between 0.9% and 2.1% of donors for each rate. Notably, the vast majority of underperforming donors were identified in only one of these categories, with fewer than 3% underperforming in more than one domain. This minimal phenotypic overlap supports the existence of biologically discrete inefficiencies and suggests that each parameter reflects a separable IVF endophenotype, likely governed by distinct molecular or cellular mechanisms. These findings highlight the inadequacy of treating IVF failure as a single outcome and underscore the value of granular phenotyping in future mechanistic investigations^3^.

Our findings demonstrate that underperformance in specific developmental phenotypes persists across multiple IVF cycles within the same donor. Among a subset of donors with three or more cycles, a statistically significant proportion exhibited repeated underperformance in the same phenotype (P < 0.001), supporting the reproducibility of these inefficiencies and suggesting they are biologically grounded rather than attributable to stochastic or technical variation. Complementing these observations, the classification model demonstrated the ability to predict outlier performance in a subsequent cycle using data from prior cycles, with consistent AUC values above 0.60 across all phenotypes. While these values fall short of the threshold typically required for clinical utility, they nonetheless reflect meaningful underlying biological signal. This supports the view that developmental inefficiencies are not random events but reflect stable, donor-specific traits, potentially underpinned by genetic or molecular mechanisms. In contrast, clinical assessment alone (i.e., without the aid of statistical modelling) is unlikely to reliably distinguish true biological outliers from background variability. This interpretation aligns with recent findings from Gill et al. (2024), that showed that even after three failed euploid blastocyst transfers, the majority of individuals went on to achieve a live birth after two more transfers, reinforcing the idea that IVF failure often reflects stochastic outcomes rather than true infertility. In the same study, the authors reported that approximately 2% of individuals undergoing multiple IVF cycles with euploid embryo transfer met criteria for biologically relevant recurrent implantation failure ^13^, a proportion that aligns well with the proportion of underperformers identified in our study for each developmental rate. Although we were unable to analyse outcomes beyond the preimplantation stage, it is possible that similar biological factors contribute to underperformance in later stages, including implantation and live birth. Together this underscores the importance of applying robust, data-driven methodologies to uncover biologically meaningful patterns in IVF outcomes and to guide future mechanistic research, even if not yet directly translatable to clinical practice.

Remarkably, our findings challenge the widely held assumption in reproductive medicine that poor IVF outcomes are inherently reflective of natural infertility. By analysing outcomes from oocyte donors (women specifically selected for their presumed fertility) we demonstrated that small but statistically significant underperformance across key IVF developmental stages can occur even in the absence of any clinical history of subfertility. This distinction underscores that IVF inefficiency does not necessarily equate to natural biological infertility. Importantly, our post-hoc survey data reinforce this interpretation: donors who underperformed in IVF did not exhibit any noticeable differences in natural fertility outcomes when compared to typical or overperforming donors. These results suggest that suboptimal IVF performance may reflect inefficiencies unique to the assisted reproduction environment, such as individual responses to ovarian stimulation or sensitivity to laboratory protocols, rather than an intrinsic reproductive defect.

However, this decoupling between natural fertility and IVF performance is likely not absolute. Notably, proven fertility, defined as documentation of a live birth prior to oocyte donation, emerged as the only measured variable with a potentially clinically relevant associated with IVF phenotypes across all developmental outcome rates. While the observed effect size was moderate and not sufficient for clinical prediction on its own, it nonetheless represents a biologically and potentially clinically relevant association. A history of successful natural conception may modestly increase the likelihood of more favourable IVF outcomes, possibly due to physiological or genetic traits that confer resilience to the stresses of ovarian stimulation or in vitro conditions. These findings suggest that proven fertility may act as a partial proxy for intrinsic reproductive robustness. However, it should not be interpreted as a definitive predictor of IVF success, but rather as one of several contributing factors to inter-donor performance variability.

Furthermore, whilst this oocyte donor cohort provides a uniquely controlled setting to study IVF endophenotypes independently of clinical infertility, infertile populations were not directly assessed. Therefore, it remains plausible, and indeed likely, that the same developmental inefficiencies observed here may be enriched or more pronounced in individuals with clinically diagnosed infertility. Supporting this, we observed a non-significant trend indicating that donors who reported taking longer than one year to conceive naturally, or who failed to conceive despite trying, were less likely to have achieved successful live births from their donated oocytes. Although this trend did not reach statistical significance, it suggests a possible continuum between subtle natural fecundity differences and IVF outcomes. These developmental endophenotypes may therefore reflect shared physiological or genetic pathways influencing both IVF efficiency and broader reproductive potential. Studying IVF underperformance in fertile individuals may therefore represents a compelling window into fundamental reproductive biology and help uncover the molecular or genetic contributors to infertility. Because the IVF process amplifies and quantifies steps that are otherwise opaque in natural conception, these endophenotypes serve as measurable proxies for gamete and embryo quality. Understanding their variation, even within fertile cohorts, informs more nuanced definitions of subfertility and allows for more individualised reproductive treatment strategies, where protocols can be tailored to specific biological profiles rather than relying on generalized treatment pathways. To fully explore the relationship between IVF inefficiency and infertility, future studies should extend similar statistical frameworks to infertile populations. Applying the same phenotyping methods will help determine whether underperformance in IVF reflects the same underlying biology in both fertile and infertile individuals, or whether the patterns observed here are unique to those who can conceive naturally. Such work will be essential for identifying which IVF endophenotypes are predictive of infertility, and which instead point to treatment-specific vulnerabilities or stochastic variation.

Importantly, our statistical modelling of IVF cycles also highlights substantial variability in single-cycle outcomes, emphasizing the role of chance or non-biological factors in IVF failure. This underscores the need for caution when interpreting poor IVF outcomes without appropriate statistical power. Although there are now several studies that have identified genes associated with gamete or embryo developmental failure, including TUBB8, PATL2, NOBOX, PADI6, TRIP13 and others, these efforts exclusively rely on arbitrarily selected outlier cases rather than a validated identification methodology ^10,14–17^. Therefore, most genetic investigations to date have focused on extreme phenotypes with complete developmental failure, often within consanguineous families, to identifying likely highly penetrant variants through case reports. While effective for detecting rare, severe defects, this strategy lacks the sensitivity to capture variants with incomplete penetrance or those contributing to more subtle, variable phenotypes. Our data-driven, methodological approach to identifying over- and underperformers in IVF helps overcome this limitation by enhancing case selection for association studies. Without such methodological rigor, there is a risk of either missing true signals or misinterpreting the effect of any associations that are uncovered when analysing more generalised datasets.

While our analysis is focused on identifying statistical underperformers, the identification of overperforming donors also holds important significance. In our dataset we identified a sum of 6.5% of donors that exhibited IVF outcomes significantly above the expected range, across one or more developmental rates. Studying overperformers can offer unique insights into the biological and procedural factors that optimize embryo development. In particular, they may help define benchmarks for ideal IVF performance and identify protective physiological traits or gene variants associated with enhanced oocyte competence, fertilization capacity, or embryo viability. Overperformers also provide a powerful contrast group for future mechanistic studies, helping to illuminate the full spectrum of reproductive efficiency and the underlying biological gradients that exist even within fertile populations. Additionally, understanding what drives exceptional IVF performance may contribute to protocol improvements that benefit all patients, regardless of fertility status.

Despite the scale and rigor of our analysis, several limitations warrant consideration. First, the study population was highly screened and selected, which likely underrepresents the full biological variability present in the general fertile population. In line with this, donors with poor outcomes in their first cycle may have been less likely to return for subsequent donations due to clinical deselection, potentially inflating the proportion of single-cycle outlier donors and biasing longitudinal analyses toward higher-performing individuals. Although we employed a multi-cycle, multi-recipient design to minimize bias, potential confounding from certain covariates, specifically recipient sperm characteristics, cannot be entirely ruled out. However, our exploratory analyses suggest that only the preimplantation development rate was statistically affected by sperm characteristics (p < 0.05), with a negligible effect size (β = 0.0003523, Kruskal-wallis test), aligning with previous findings that the sperm play a limited role in IVF outcomes when using donor oocytes^18^. Finally, while our statistical framework is effective at identifying performance outliers, it does not elucidate the underlying biological mechanisms driving exceptionally poor or exceptionally good outcomes. A critical next step will be to integrate this phenotyping model with genomic, transcriptomic, or proteomic data to uncover molecular drivers of IVF inefficiency and success, which could form the basis for future functional studies.

In conclusion, this unprecedented study demonstrates that poor IVF outcomes can occur independently of infertility, even among rigorously screened, naturally fertile oocyte donors. By statistically defining underperformance across key stages of the IVF process, we establish a robust framework for phenotyping IVF inefficiency and lay the groundwork for future research into its biological underpinnings. These findings challenge conventional assumptions, emphasize the need for individualized IVF strategies, and open new avenues for precision reproductive medicine.

## Methods

### Study design and overview

This study is a retrospective cohort analysis of oocyte donation IVF cycles conducted at IVIRMA Spain between 1999 and 2023, with a prospective follow-up component assessing natural fertility outcomes in a selected donor population. All donors were recruited in accordance with Spanish national legislation (Law 14/2006). Donors were healthy women aged 18–35 years who passed a comprehensive fertility assessment as well as general medical, psychological and genetic screening to ensure suitability for oocyte donation. Matching of donors to recipients was carried out by clinical staff based on phenotypic characteristics.

The primary objectives were to establish a baseline for IVF over and underperformance in a population of presumed fertile individuals and evaluate the prevalence of extreme IVF outcomes that cannot be explained by the role of chance. The dataset comprises anonymized records from 32.492 oocyte donors who underwent 84.228 oocyte retrievals, resulting in 114.237 IVF cycles for oocyte recipients. All oocyte donors included in this study provided informed consent at the time of donation, agreeing to the use of their clinical data for research purposes and to being recontacted for follow-up studies. The study received ethical approval from the ethics review board at Centro de Investigación de IVI under approval reference number 2304-JUN-052-AC and all procedures adhered to the ethical guidelines set forth by the Declaration of Helsinki and the regulatory requirements governing human reproductive research in Spain. In addition, oocyte donors participating in the prospective survey on natural fertility provided separate informed consent before completing the questionnaire and were fully informed about the study objectives, data confidentiality measures, and their right to withdraw at any time.

### Ovarian stimulation and IVF procedures

Ovarian stimulation, IVF, and embryo culture were performed using standard, validated protocols at each participating clinic. Following ovulation trigger, oocyte retrieval was performed, and the number of cumulus–oocyte complexes (COCs) retrieved was recorded. Oocytes were assessed for maturity after cumulus cell removal, with mature (metaphase II; MII) oocytes identified by the presence of the first polar body.

Insemination was performed using either standard IVF or intracytoplasmic sperm injection (ICSI) techniques. Fertilization was assessed and recorded based on the presence of two pronuclei (2PN). Resulting 2PN embryos were cultured in vitro, and the number of clinically usable blastocysts was recorded.

Due to the longitudinal nature of the dataset and the extended period over which the oocyte donation cycles were performed, there were significant changes to clinical protocols over time. These included the introduction and increasing use of vitrification techniques, the adoption of gonadotrophin releasing hormone (GnRH) antagonist protocols for ovarian stimulation allowing triggering with either GnRH agonist or human chorionic gonadotrophin (hCG), and a shift from embryo utilisation at the cleavage stage to almost exclusive blastocyst culture (Supplementary figure 2). Such changes have known impacts on IVF outcomes. To account for these temporal variations, changes in protocols and practices were systematically measured and statistically controlled for in the analysis.

### Outcome measures and statistical analysis plan

To systematically characterize IVF performance, we defined four key developmental rates reflecting distinct stages of the IVF process and potentially different biological processes: 1) Oocyte Maturation Rate – Defined as the proportion of retrieved oocytes that reached the MII stage, providing a measure of oocyte competence for fertilization. 2) Fertilization Rate – Defined as the proportion of MII oocytes that successfully formed 2PN zygotes following exposure to sperm. 3) Embryo Utilization Rate – Defined as the proportion of fertilized oocytes (2PN zygotes) that developed into embryos of sufficient quality to be transferred or cryopreserved. 4) Preimplantation Development Efficiency – A composite phenotype integrating the maturation, fertilization, and embryo utilization rates to assess the overall efficiency of preimplantation development. This metric accounts for cumulative losses at each stage, with the outcome being blastocyst formation relative to the total number of COCs retrieved. These rates allow for a structured analysis of IVF inefficiencies by distinguishing specific points of failure, thereby enabling a more refined understanding of IVF underperformance in the presumed fertile donor oocyte population.

QC steps involved removing records with missing or inconsistent values that could compromise the accuracy of rate calculations. This included verifying correct data input by ensuring the number of oocytes, zygotes, and blastocysts recorded aligned with expected biological and procedural constraints. Additionally, the preimplantation development efficiency rate was only computed for donors who donated all retrieved oocytes in each cycle to ensure data completeness.

To assess selection bias after QC, the distributions of demographic variables and other parameters of interest before and after QC were compared by applying the Wilcoxon Signed-rank test (for continuous variables) and the Chi-Square Goodness of Fit test (for categorical variables). Given the large sample size, we computed effect sizes with Cliff’s Delta *d* (for continuous variables) and Cramer’s *V* (for categorical variables) to quantify the variability due to real biological differences. Effects were considered as negligible with |*d|* < 0.2 or *V* < 0.1, small with 0.2 < |*d|* < 0.5 or 0.1 < *V* < 0.3, medium with 0.5 < |*d|* < 0.8 or 0.3 < *V* < 0.5, large otherwise^21,22^.

Furthermore, a post-QC univariate analysis was conducted to explore associations between all available variables and each developmental rate outcome. Spearman correlation, the Wilcoxon signed-rank test, and the Kruskal-Wallis test were applied for continuous, binary, and categorical variables, respectively (see Supplementary Table 4), to identify any key variables that required adjustment in the subsequent statistical modelling.

Categorisation of donors as statistical over and underperformers in each developmental rate was performed by employing a combination of Binomial tests and Mixed Effect models^23^. The Binomial test treated each donor’s cumulative outcomes across cycles as the result of a Bernoulli trial, comparing observed rates to expected probabilities under the null hypothesis, with statistical significance adjusted using Bonferroni correction. Mixed Effect models utilized Binomial Regression with a logit link function to estimate performance while accounting for confounding factors including donor age, year of donation, number of stimulation cycles, number of follicles identified during monitoring scans and the type of trigger used (namely hCG, GnRH-a or both). Donor ID was included as a random effect to control for repeated stimulations within individuals.

Donors were classified as statistically underperforming or overperforming with Binomial-derived P<0.05 and if the rate predicted by the Mixed Effect models deviated from the mean by more than two standard deviations. A donor was included in the final list of over and underperformers if flagged by at least one of the statistical approaches, ensuring an increased power for potential procedural or genetic association studies. Finally, to confirm the accuracy of the selection, an expert-based inspection of the results was performed before implementing further statistical analyses.

The distributions of demographic variables and other parameters of interest within different categories of donors (i.e., underperforming, overperforming, average) were compared by applying the Kruskal-wallis (for continuous variables) and the Chi-Square Goodness of Fit test (for categorical variables). To account for the unbalanced sample sizes between the three groups of donors, we computed effect sizes with Epsilon Squared ε (for continuous variables) and Cramer’s V (for categorical variables) that represent the proportion of the variance due to true group differences. Post-hoc tests were applied to evaluate the differences between each pair of groups (Conover’s test and Chi-Square post-hoc test for continuous and categorical variables, respectively). Effects were considered as negligible with ε < 0.01 or *V* < 0.1, small with 0.01 < ε < 0.06 or 0.1 < *V* < 0.3, medium with 0.06 < ε < 0.14 or 0.3 < *V* < 0.5, large with ε > 0.14 or *V* >= 0.5^24^.

For each rate, the statistical impact of the sperm sample on donor performances was assessed via Kruskal-wallis test for sperm source and with a Beta regression model for sperm concentration.

To evaluate whether the occurrence of consecutive cycles with significantly poor or good performance could be attributed to chance, we focused on donors with three or more stimulation cycles and applied a Binomial test, using the expected rate derived from the overall distribution of outcomes across all cycles. In parallel, using the same data subset, we assessed our model’s ability to identify statistical outliers across successive treatment cycles from the same donor by developing a supervised classification model based on a Logistic Regression algorithm. For each of the four developmental rates, the model was trained to predict whether a donor would be a statistical over- or under-performer in the next stimulation cycle, using data from the immediately preceding cycle as input. Predictive performance was evaluated using receiver operating characteristic (ROC) curves and the corresponding area under the curve (AUC) values. Together, these complementary approaches provide a framework for distinguishing biologically meaningful patterns from random fluctuation or technical variability in IVF treatment outcomes.

Statistical analyses were performed using custom scripts in Python (version 3.10.8) and R (version 4.3.1).

### Survey of natural fertility post oocyte donation

A structured questionnaire was developed to assess the natural fertility of oocyte donors, focusing on key metrics such as history of natural conception, time to pregnancy, reproductive success, and adverse pregnancy outcomes including miscarriage and foetal or obstetrical complications (Supplementary Figure 3). The questionnaire was designed to minimize recall bias through standardized formats and response options. Donors who underwent ovarian stimulation between 2000 and 2015 and were identified as statistical overperformers or underperformers were selected for participation. A randomly selected group of typical performers served as a control cohort. A subset of donors identified as statistical outliers in either embryo utilisation or preimplantation development rates, considered the most relevant proxies for natural fecundity, were further analysed for pregnancy outcomes from their donation cycles, to explore more in-depth potential associations with their questionnaire responses. Donors were contacted by phone and invited to participate, and the questionnaire was administered by experienced IVF clinicians. Only fully completed questionnaires were included in the analysis, and all responses were anonymized. Statistical comparisons across donor performance categories were performed using chi-squared or Fisher’s exact tests, with response categories aggregated where needed to ensure adequate statistical power.

## Data Availability

All data produced in the present study are available upon reasonable request to collaborate with the authors

