## Supplementary figures and tables for "Mapping performance in 114,237 IVF cycles with donor oocytes reveals reproducible inefficiencies and distinct endophenotypes"

**Supplementary figure**

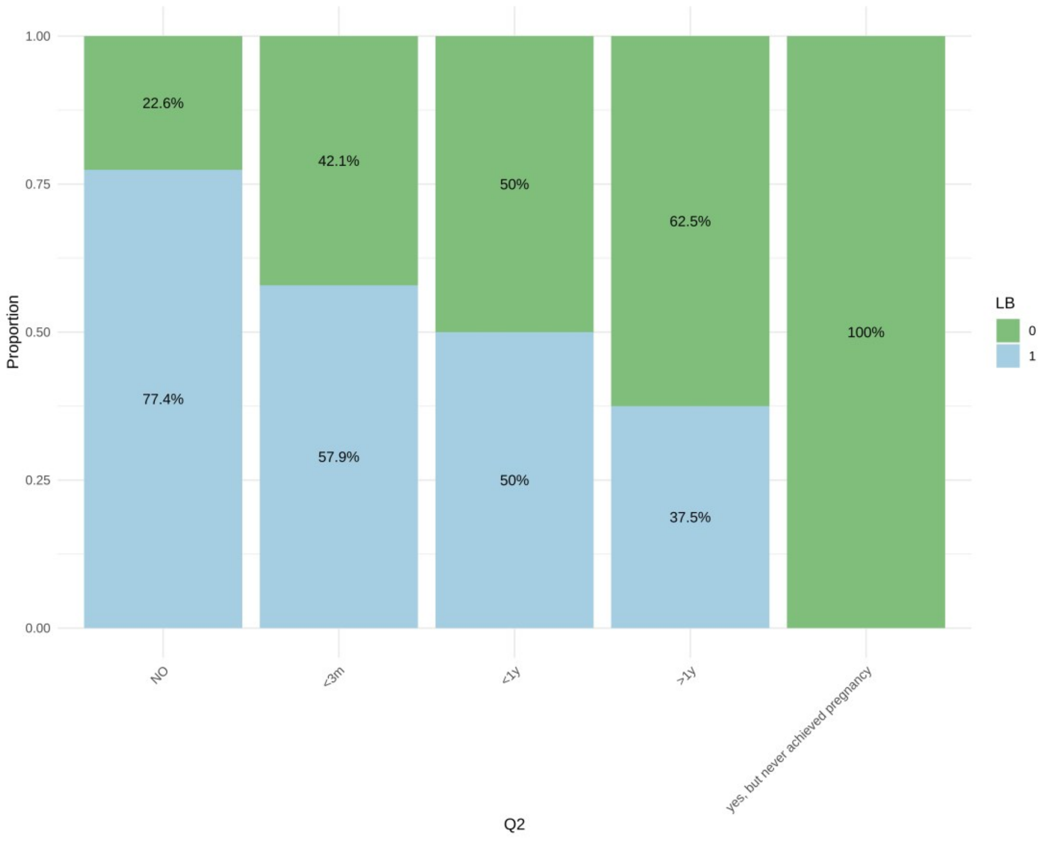


***Supplementary Figure 1. Responses to Question 2 of the questionnaire in the donor sub-cohort with known IVF live birth outcomes.*** *This figure displays the distribution of donors whose donated oocytes resulted in at least one live birth (blue) versus those with no live birth outcome (green), based on their responses to Question 2: the time it took to conceive naturally after donation (if applicable). The five response categories reflect increasing time to conception, including an option for unsuccessful attempts.*

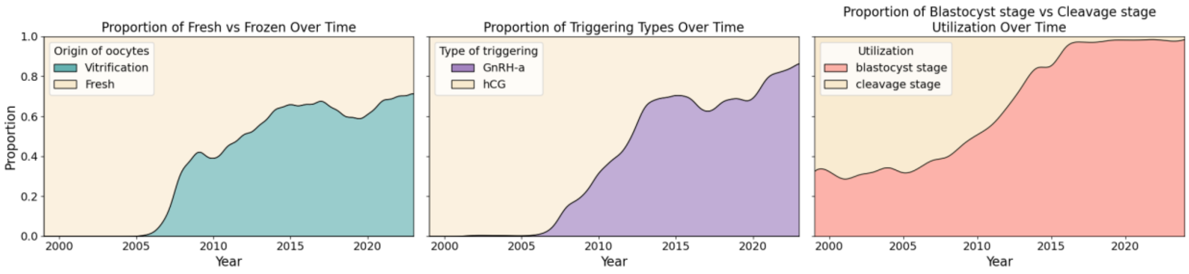


***Supplementary figure 2. Clinical utilisation of vitrification, GnRH agonist triggering and blastocyst culture over the study period.*** *Density plots representing the proportion of each procedure over the dataset period.*

**Project Code** **Questionnaire Natural Fertility Assessment**

**NHC egg donor:**
**Name of physician:**
**Date of data collection:**

**Q1: Did you give birth to a child conceived naturally prior to egg donation?**
- ☐ Yes
- ☐ No

**Q2: Since donation have you tried to conceive naturally (without medication)?**
- ☐ Yes, fell pregnant within 3 months of trying to conceive
- ☐ Yes, fell pregnant within 1 year of trying to conceive
- ☐ Yes, fell pregnant after over a year of trying to conceive
- ☐ Yes, but never achieved a pregnancy
- ☐ No

*If you replied No to the previous question:*

**Q3: Did you get IVF treatment?**
- ☐ Yes
- ☐ No

*If you replied Yes to the previous question:*

**Q4: Did your IVF treatment end up in a live birth?**
- ☐ Yes
- ☐ No

*If you conceived naturally after egg donation:*

**Q5: How many natural pregnancies did you achieve after egg donation?**
- ☐ None
- ☐ One or more than one
  Please specify the number: ___

*If you replied One or more to the previous question:*

**Q6: Were any of your pregnancies with multiple gestations (i.e., twins, triplets) after egg donation?**
- ☐ No
- ☐ Yes, twins
- ☐ Yes, triplets

**Q7: How many pregnancy losses did you experience after egg donation?**
- ☐ None
- ☐ One or more than one
  Please specify the number: ___

**Q8: How many live births did you achieve after egg donation?**
- ☐ None
- ☐ One or more than one
  Please specify the number: ___

*If you replied One or more than one to the previous question:*

**Q9: Were any of your babies born with congenital abnormalities after egg donation?**
- ☐ No
  Specify the number of healthy new-borns: ___
- ☐ Yes
  Specify the number of babies with congenital abnormalities: ______
  Please specify the congenital abnormality: ______________________

______________________________________________________________________
______________________________________________________________________
______________________________________________________________________
______________________________________________________________________

**Q10: Have any of your children conceived after egg donation been diagnosed with a genetic disorder or neurodevelopmental delay/disorder?**
- ☐ No
- ☐ Yes
  Specify the number of affected children: ___
  Please specify the diagnosis: ________________________________

______________________________________________________________________
______________________________________________________________________
______________________________________________________________________
______________________________________________________________________

***Supplementary Figure 3. Questionnaire distributed to subset of selected donors to assess their natural fertility post oocyte donation.***

### Supplementary tables

**Supplementary Table 1:**

| Variable | Test/Model | Fertilization | | Embryo-Utilization | | Pre-implantation | |
|---|---|---|---|---|---|---|---|
| | | P-value | Effect size | P-value | Effect size | P-value | Effect size |
| Sperm Source | Kruskal-Wallis test | 1 | n/a | 0.026 | n/a | 0.116 | n/a |
| Sperm Origin | Kruskal-Wallis test | 1 | n/a | 0.167 | n/a | 0.197 | n/a |
| Sperm Concentration | Beta Model | 0.063 | n/a | 0.171 | n/a | 0.0004 | negl. |

***Supplementary Table 1. Sperm-related factors and their impact on fertilization or embryo development outcomes in oocyte donation cycles.*** *The analysis looked at sperm source (e.g., partner or donor, ejaculated or surgically retrieved) and sperm concentration. P-values show whether there were statistically significant differences between the QC subsets and the full dataset. Effect sizes indicate how large those differences were: "negl." means negligible (likely not meaningful), "n/a" means not applicable (the difference was not statistically significant, so no effect size was calculated).*

**Supplementary Table 2:**

| Question | Fertilization | Maturation | Embryo-Utilization | Pre-implantation |
|---|---|---|---|---|
| | P-value | P-value | P-value | P-value |
| $Q_1$ | 0.585 | 0.711 | 0.049 | 0.812 |
| $Q_2$ | 0.776 | 0.948 | 0.336 | 0.326 |
| $Q_3$ | 1 | 0.456 | 0.081 | 0.211 |
| $Q_4$ | 1 | 0.320 | 0.135 | 0.178 |
| $Q_5$ | 1 | 1 | 1 | 1 |
| $Q_6$ | 1 | 1 | 0.724 | 1 |
| $Q_7$ | 1 | 0.808 | 0.352 | 0.922 |
| $Q_8$ | 1 | 0.813 | 0.777 | 0.027 |
| $Q_9$ | 1 | 1 | 1 | 1 |
| $Q_{10}$ | 1 | 1 | 0.290 | 1 |

***Supplementary Table 2. Analysis of differences in questionnaire responses across donor performance categories for each developmental rate.*** *This table summarizes the differences between self-reported questionnaire responses among donors classified as underperformers and typical performers plus overperformers across the four IVF developmental rates. Statistical comparisons were conducted to assess whether natural fertility indicators differed by IVF performance category.*

**Supplementary Table 3:**

| Question | Live Birth | Miscarriage |
|---|---|---|
| | P-value | P-value |
| $Q_1$ | 0.644 | 1 |
| $Q_2$ | 0.0436 | 0.605 |
| $Q_3$ | 1 | 1 |
| $Q_4$ | 1 | 0.621 |
| $Q_5$ | 0.398 | 0.336 |
| $Q_6$ | 1 | 1 |
| $Q_7$ | 0.7113 | 1 |
| $Q_8$ | 0.088 | 0.061 |

***Supplementary Table 3. Analysis of differences in live birth and miscarriage outcomes across questionnaire responses in the outlier donor subgroup.*** *This table presents statistical comparisons of live birth and miscarriage rates among donors classified as outliers in embryo utilisation or preimplantation development, for whom pregnancy outcome data were available. Differences were assessed for each relevant questionnaire item to evaluate potential associations with natural fertility indicators.*

**Supplementary Table 4:**

| Variable | Maturation | | | Fertilization | | | Embryo-Utilization | | | Pre-implantation | | |
|---|---|---|---|---|---|---|---|---|---|---|---|---|
| | P-value | Eff. size | Magn. | P-value | Eff. size | Magn. | P-value | Eff. size | Magn. | P-value | Eff. size | Magn. |
| Age | <0.001 | 0.029 | negl. | 0.081 | n/a | n/a | <0.001 | 0.040 | negl. | <0.001 | 0.045 | negl. |
| BMI | 0.836 | n/a | n/a | 0.014 | n/a | n/a | 0.841 | n/a | n/a | 0.157 | n/a | n/a |
| Stimulation Dose | 0.005 | 0.019 | negl. | 0.141 | n/a | n/a | 0.153 | n/a | n/a | 0.015 | n/a | n/a |
| Days of Stimulation | <0.001 | 0.047 | negl. | <0.001 | -0.008 | negl. | 0.073 | n/a | n/a | 0.781 | n/a | n/a |
| E2DHCG | 0.128 | n/a | n/a | 0.325 | n/a | n/a | <0.001 | -0.011 | negl. | 0.123 | n/a | n/a |
| P4DHCG | 0.258 | n/a | n/a | 0.863 | n/a | n/a | <0.001 | -0.061 | negl. | <0.01 | -0.052 | negl. |
| Year | <0.001 | -0.01 | negl. | <0.001 | 0.114 | small | 0.2241 | n/a | n/a | <0.001 | 0.021 | negl. |
| Clinica | <0.001 | 0.011 | small | <0.001 | 0.008 | negl. | <0.001 | 0.023 | small | <0.001 | 0.019 | small |
| Doctor | <0.001 | 0.015 | small | <0.001 | 0.016 | small | <0.001 | 0.022 | small | <0.001 | 0.016 | small |
| Ethnic bakground | <0.001 | 0.000 | negl. | 0.304 | n/a | n/a | <0.001 | 0.000 | negl. | 0.522 | n/a | n/a |
| Type of triggering | 0.6447 | n/a | n/a | <0.001 | 0.024 | small | <0.001 | 0.000 | negl. | 0.657 | n/a | n/a |
| Origin/status oocytes | n/a | n/a | n/a | <0.001 | -0.077 | negl. | <0.001 | 0.111 | negl. | <0.001 | 0.203 | small |
| OCP | <0.001 | -0.043 | negl. | <0.001 | -0.050 | negl. | 0.002 | 0.020 | negl. | 0.449 | n/a | n/a |
| Type of protocol | <0.001 | 0.000 | negl. | <0.001 | 0.027 | small | <0.001 | 0.000 | negl. | <0.001 | 0.002 | negl. |

***Supplementary Table 4. Univariate analysis of all available variables with the outcomes of each developmental rate.*** *P-values show whether there were statistically significant relationships between each rate and each variable. Effect sizes quantify how large those differences were, as shown by Magnitude levels, where "negl." means negligible (likely not meaningful), "n/a" means not applicable (the relationship was not statistically significant, so no effect size was calculated).*
